# Regional variation of cancer incidence in Panama

**DOI:** 10.1101/2025.03.17.25324028

**Authors:** María Moya-Navamuel, Timothy M Thomson

**Author notes:** <u>Correspondence</u>: Timothy M Thomson, INDICASAT-AIP, Panama. <u>Funding</u>: none.

## Abstract

The variability of cancer incidence between diverse geographical locations has been attributed to a broad range of factors, including environmental and genetic. The geographical co-localization of cancer incidence and environmental factors is a valid initial approach to test causality hypotheses. Herein, we have analyzed the incidence of major cancer types in the Provinces and Comarcas of Panama, from 2018 through 2022. We have found a striking regional variability, with significantly higher incidence of colorectal, breast and prostate cancers in the Azuero Peninsula as compared to other regions or to the total national incidence. These observations warrant a comprehensive analysis of environmental toxicants in the entire territory of Panama, so as to properly address causal hypotheses that may explain the significant regional variation in cancer incidence.

## Introduction

Neoplastic tumors arise as clusters of cells in which growth regulation and integration into tissue patterns are altered through genetic, epigenetic or metabolic mechanisms [1, 2] or through interactions with microorganisms [3, 4], and subsequently escape immune surveillance [5]. There are many recognized endogenous and exogenous drivers of initial neoplastic events and subsequent immune escape, which operate against a backdrop of idiosyncratic scenarios, including genetic susceptibility [6]. Because of the vast geographical diversity of genetic and environmental factors and their multiple combinations that may be linked to neoplasia initiation and progression, large disparities are observed in the incidence of neoplastic diseases between different regions worldwide [7], and within a single region or country [8, 9].

Analyses of global neoplastic disease burden suggest that local or regional environmental factors play a significant role in the emergence of many common neoplasms, with genetic factors playing a more prominent role in the emergence of specific types and subtypes of cancers [10, 11]. A classic example of the likely impact of environmental factors in cancer incidence is that of prostate cancer among Japanese men who immigrated to the United States, with significantly higher incidence than for Japanese men living in Japan [12]. The differences in cancer incidence between immigrants and populations that remain in their homelands is generally attributed to changes in lifestyle, diet and additional environmental factors [13]. More generally, exposure to environmental chemical agents (e.g., herbicides), physical agents (e.g., radiation) or infectious agents (e.g., papillomavirus, hepatitis B virus or *Helicobacter pylori*) has been linked to the variable geographical incidence of numerous cancers [14-16], with suggestions of causal mechanisms including direct mutagenicity [17], reassortment of the gut microbiota [3], with direct and indirect effects on the risk of developing cancers [18, 19], or modulation of the immune system resulting in debilitated immune surveillance [20]. As such, geolocalization of cancers, in conjunction with geolocalization of specific environmental factors, is a valid approach to unveil potential causal agents [10, 21], which should be followed by mechanistic studies and the adoption of preventative public health measures.

Herein, we have analyzed the regional incidence of common cancers in Panama, using data available through the National Cancer Registry of the Ministry of Health (www.minsa.gob.pa), from 2018 to 2022. We have found significant provincial variations in the incidence of most cancers in the database. With the exception of skin and prostate cancers, we find a trend towards increased incidence, nationwide, of all cancers during this 5-year period, with a sharp decline in 2020 and return to augmented incidence in subsequent years, which we attribute to the impact of the COVID-19 pandemic in cancer detection and diagnosis. Notably, two provinces in the Azuero peninsula, namely Herrera and Los Santos, present significantly higher incidence, as compared to the overall national incidence, of gastric, colorectal, lung, skin, breast and prostate cancers. The evidence provided here warrants a search for potential causal factors that may be mitigated by targeted public health interventions.

## Results

In order to determine the variations in cancer incidence among Provinces and indigenous territories (Comarcas) in Panama, we resorted to the National Cancer Registry (NCR) of the Ministry of Health, which maintains a registry of the most common types of cancer, classified by anatomical location and Province or Comarca, from 2018 through 2022. The types of cancers at the NCR include gastric, colorectal, lung, skin, breast, cervical and prostate cancers, without specifications of subtypes. The incidence per 100,000 inhabitants was calculated based on the case numbers in the NCR and the official 2020 census, as available through the National Institute of Statistics and Census (Instituto Nacional de Estadística y Censo, INEC), and population estimates for other years calculated by the Mapa de Información Económica de la República de Panamá (MINERPA) of the Interamerican Development Bank (Banco Interamericano de Desarrollo, BID).

We observed an overall upward trend in incidence for all cancer types from 2018 to 2022, with the exception of a decline in 2020, particularly sharp for skin, breast, cervical and prostate cancers, but also evident for gastric and colorectal cancers (Figure 1, Supplemental Figure 1). Most Provinces and Comarcas followed this pattern for the majority of tumor types, with few exceptions. For example, the incidence did not decline in 2020 for gastric cancer in Herrera or Panamá Oeste, or for colorectal cancer in Veraguas (Figure 1). The sharp decline in cancer incidence in 2020 may be attributable to the generalized reduction in apparent disease burden during the peak of the COVID-19 pandemic, owed to the stalling of health systems, including cancer diagnosis and registry [22]. Remarkably, after 2020, the incidence of various cancers showed strong increases in many regions, above pre-2020 pandemic peak levels (Figures 1 and 2). The only cancer type that did not follow an upward trend after 2020 were skin cancers, which showed a decline in incidence after the COVID-19 pandemic peak in several regions, most pronounced in Herrera (Figures 1, 2 and Supplemental Figure 1). The Comarcas, autonomous regions inhabited by indigenous peoples, had sparse data and the apparent incidence for all cancer types was lower than that of all other regions. We attribute the data scarcity in the Comarcas to under-registry due to deficiencies in the healthcare and cancer registry systems, rather than *bona fide* lower incidences. Consequently, we excluded the Comarcas from statistical analyses (Figure 2).

**Figure 1.**
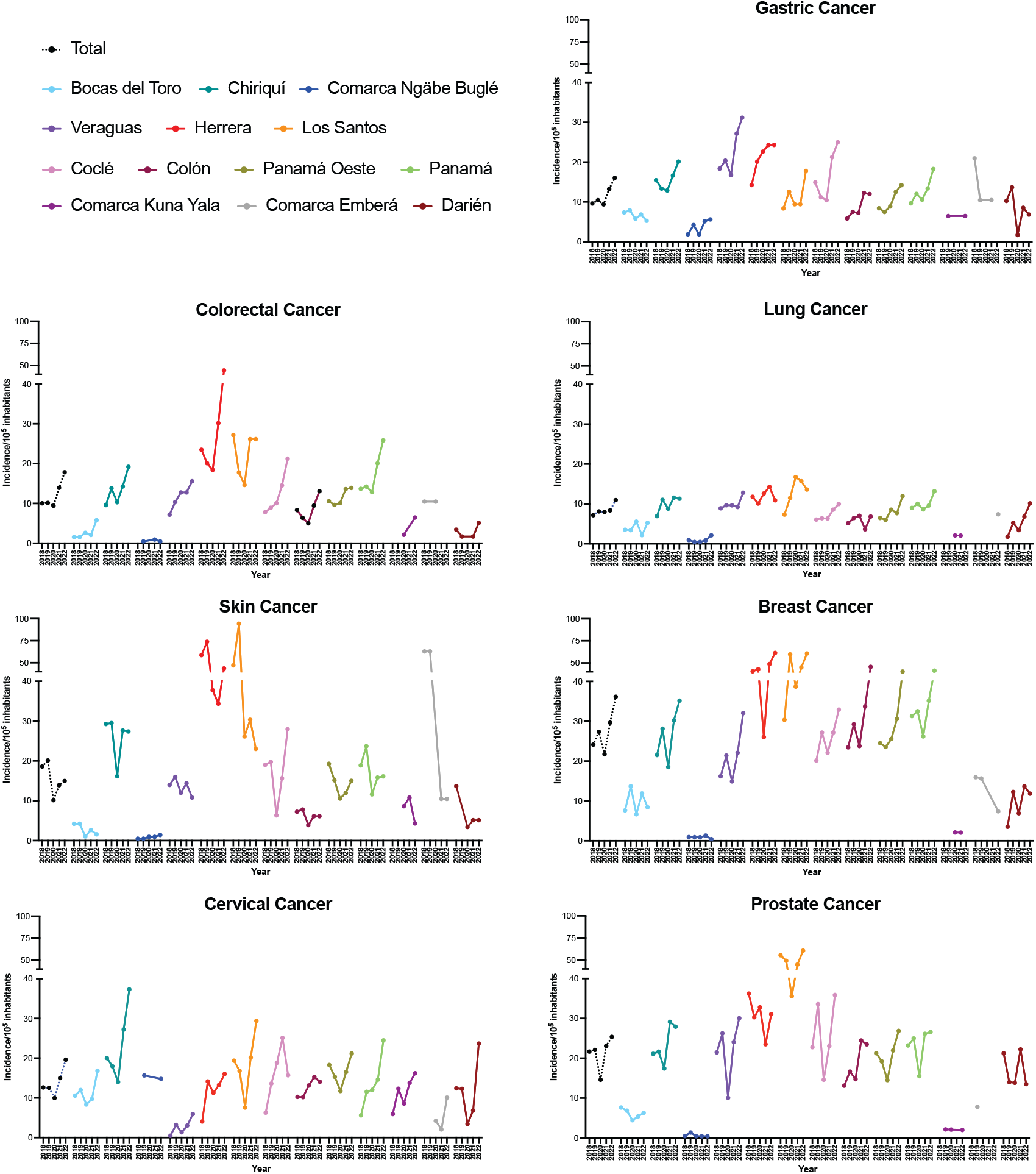
Incidence of cancer cases per 100,000 inhabitants, per Province or Comarca for the years 2018 through 2022. Case numbers for each cancer type were accessed through the National Cancer Registry of the Ministry of Health (https://www.minsa.gob.pa/contenido/registro-nacional-del-cancer), and normalized by population censed by the Instituto Nacional de Estadística y Censo (inec.gob.pa) or population estimated by the Mapa de Informació n Econó mica de la República de Panamá (MINERPA), of the Interamerican Development Bank (https://minerpa.com.pa/poblacion-por-provincia-y-genero/).

**Figure 2.**
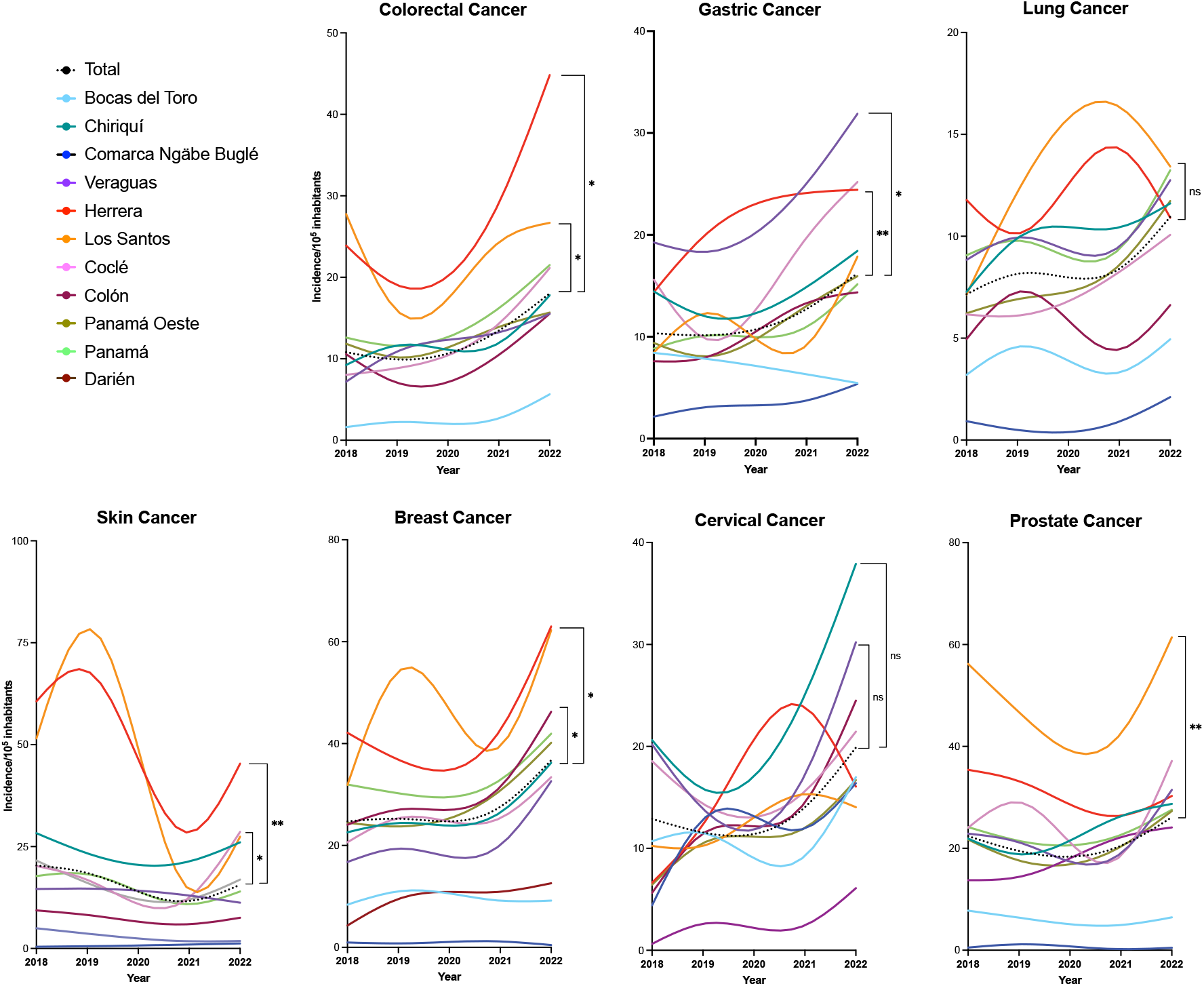
Between-Province comparisons of cancer-specific incidence. For each cancer type, population-normalized incidence was compared to the total national incidence (dotted lines) and analyzed for significance by Welch’s t-test. Fitted curves were generated by applying spline/LOWESS to longitudinal data. * denotes *p* ≤ 0.01; ** denotes *p* ≤ 0.001; ns = not significant.

Two Provinces, Herrera and Los Santos, stood out above all other Provinces in that they presented incidences of several types of cancer that were significantly higher than the total national incidence (Figure 2). This was particularly evident for colorectal, breast and prostate cancers (Figure 2). In 2022, the incidence per 10^5^ inhabitants in Herrera was 44.51 for colorectal, 61.31 for breast and 31.07 for prostate cancers. In Los Santos, it was 26.17 for colorectal, 60.71 for breast and 60.71 for prostate cancers (Supplemental Table 1). In comparison, in 2022, the countries with the highest age-standardized rates in the world for colorectal cancer was Denmark (48.07), for breast cancer, France (105.42) and for prostate cancer, Lithuania (135.04) (GLOBOCAN, gco.iarc.fr). In Latin America, the countries with the highest incidence for colorectal cancer in 2022 were Uruguay (31.02) and Argentina (24.2); for breast cancer, Uruguay (75.05), Argentina (73.02) and Brazil (63.12); and for prostate cancer, Brazil (76.3), Uruguay (58.96) and Cuba (58.11). The incidence of gastric cancer was also significantly higher in Herrera than in most other regions and also compared to national incidence, until 2022, a year in which the incidence in Veraguas, 31.26 per 10^5^ inhabitants, surpassed that of Herrera, 24.35 per 10^5^ inhabitants (Supplemental Table 1). In 2022, the country with the highest incidence in the world for gastric cancer was Mongolia (35.49 per 10^5^ inhabitants) and, in Latin America, Peru (14.31) and Chile (14.18). Therefore, the incidence for colorectal, breast, prostate and gastric cancers in Herrera or Los Santos is at least comparable to that of some of the highest incidence Latin American countries.

An exception to the general higher cancer incidence in Herrera and Los Santos was cervical cancer, the highest incidence being in Chiriquí (37.31 per 10^5^ inhabitants in 2022), although without reaching statistical significance when compared to the national incidence (Figure 2). Importantly, Herrera and Los Santos are contiguous Provinces that occupy most of the Azuero Peninsula (Supplemental Figure 1). The remaining area of Azuero is occupied by Veraguas. Therefore, the Azuero Peninsula harbors the highest incidence of major cancer types in Panama.

## Discussion

Geographical disparities in cancer incidence may suggest exposure to environmental carcinogenic factors, which may be unevenly distributed at different locations. The contemporaneous occurrence of increased cancer incidence and a specific event, such as the acute exposure of the population of a specific site to large doses of a known or suspected carcinogen, renders the environmental carcinogen hypothesis particularly strong [23, 24]. In the absence of such acute events, the attribution of excess cancer incidence in a given region to specific environmental carcinogens requires additional evidence that substantiates causality. There are numerous studies associating various cancers to chronic exposure to environmental agents, including smoke [25], pesticides and herbicides [26], heavy metals [27], various forms of radiation [28], including sunlight [29], or diet [18, 30]. Other studies have found associations of cancer incidence to socioeconomic determinants, such as extreme poverty [31], a category that encompasses many of the factors mentioned above.

We have found a significantly higher incidence of major cancer types in a specific region in Panama, the Azuero Peninsula, as compared to the rest of the country, sustained over time. Azuero is a predominantly agricultural region, not characterized by a socioeconomic underprivileged population (minerpa.com.pa), and with relatively low representation of afrodescendant or indigenous populations (Instituto Nacional de Estadística y Censo, inec.gob.pa). This suggests that socioeconomic determinants may not be likely major factors for consideration in causal attribution. On the other hand, corn is the largest crop in Herrera and Los Santos, which together produce close to 90% of all corn in Panama (Ministerio de Desarrollo Agropecuario, mida.gob.pa). At least 10 different herbicides are used in corn farming in Panama, including atrazine, pendimethalin, nicosulfuron or glyphosate (Instituto de Innovación Agropecuaria de Panamá, idiap.gob.pa). Relevantly, high levels of atrazine have been detected in Azuero following an acute spillover event (Ministerio de Salud, minsa.gob.pa). However, no nationwide survey of herbicide levels or exposure has been conducted to date. Atrazine, banned in the European Union [32], is the most commonly detected pesticide contaminating drinking water in the United States [33]. Atrazine is an endocrine disruptor [34] and an immunomodulator [35], and its exposure has been associated with an increased incidence of several types of cancer [33].

Our findings warrant a comprehensive analysis of environmental toxicants in Panama, particularly in the Azuero Peninsula, in order to address the hypothesis that exposure to specific compounds may be causally linked to the very high incidence of cancer in that region.

## Methods

### Data sources

Data for cancer incidence in Panama, stratified by Province or Comarca and year, were accessed through the National Cancer Registry of the Ministry of Health (https://www.minsa.gob.pa/contenido/registro-nacional-del-cancer). The accessible data include case numbers for gastric, colorectal, lung, skin, breast, cervical and prostate cancers, with no indication of histological or molecular subtypes within these anatomical locations, stratified by year, Province, Comarca, sex and age range. Demographic and population census data were accessed through the Instituto Nacional de Estadística y Censo (inec.gob.pa). Population estimates for non-census years were obtained from the Mapa de Información Económica de la República de Panamá (MINERPA), of the Interamerican Development Bank (https://minerpa.com.pa/poblacion-por-provincia-y-genero/).

### Geolocalization and map rendering

Cancer incidence data were imported into ESRI ArcGIS Pro 3.4. Spatial data were incorporated by uploading the “Panama Province Boundaries 2024” layer from the ArcGIS Online Portal [36], which contains polygon features for administrative divisions. Using the “Add Join” function, the cancer incidence dataset was linked to the polygon layer based on the “Province Name” attribute. The final joined dataset enabled spatial visualization, with incidence values mapped using the “Unclassed Colors” symbology on the cancer incidence field to represent variations across Provinces or Comarcas for each cancer type and year.

### Statistical analysis

Longitudinal data were fitted by applying a cubic spline/LOWESS. For statistical comparisons, Welch’s t-test was applied for paired samples assuming non-Gaussian distribution.

## Data Availability

All data produced in the present work are contained in the manuscript

## Acknowledgments

We thank Emilce Mejía and Alejandra Rivera, of ESRI Panamá, for assistance in geolocalization analyses, and Patricia Llanes Fernández and Edgardo Castro, of INDICASAT, for helpful discussions.

## Funding

This study did not receive any specific funding.

## Author contributions

MM-N performed analyses and bibliographical searches, generated figures and edited the manuscript. TMT designed the study, performed analyses, generated figures and wrote the manuscript.

## Conflicts of interest declaration

The authors declare no conflicts of interest.

## Notes

<u>Conflicts of interest</u>: none

### Competing Interest Statement

The authors have declared no competing interest.

### Funding Statement

This study did not receive any funding

### Author Declarations

The study used ONLY openly available human data that were originally located at: https://www.minsa.gob.pa/contenido/registro-nacional-del-cancerhttps://minerpa.com.pa/ https://gco.iarc.fr/en

