## Supplemental Figure 1 for "Regional variation of cancer incidence in Panama"

### Supplemental Files

**Supplemental Figure 1**. Geolocalization of cancer incidence in the Provinces and Comarcas of Panama from 2018 through 2022. For each cancer type, colors are graded from 0 to the highest provincial incidence in the entire 5-year period. Uncolored areas denote absence of registry data.

**Gastric Cancer**

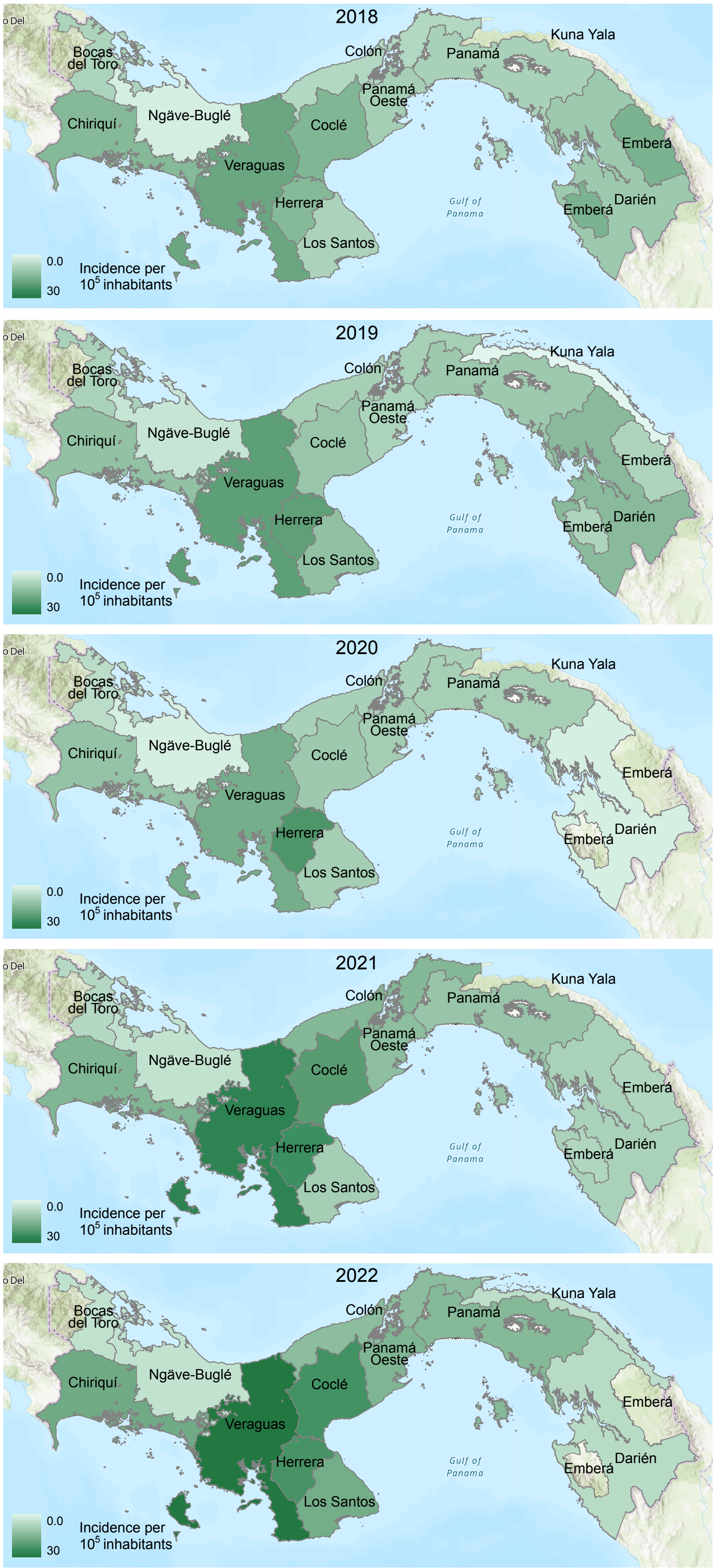

### Colorectal Cancer

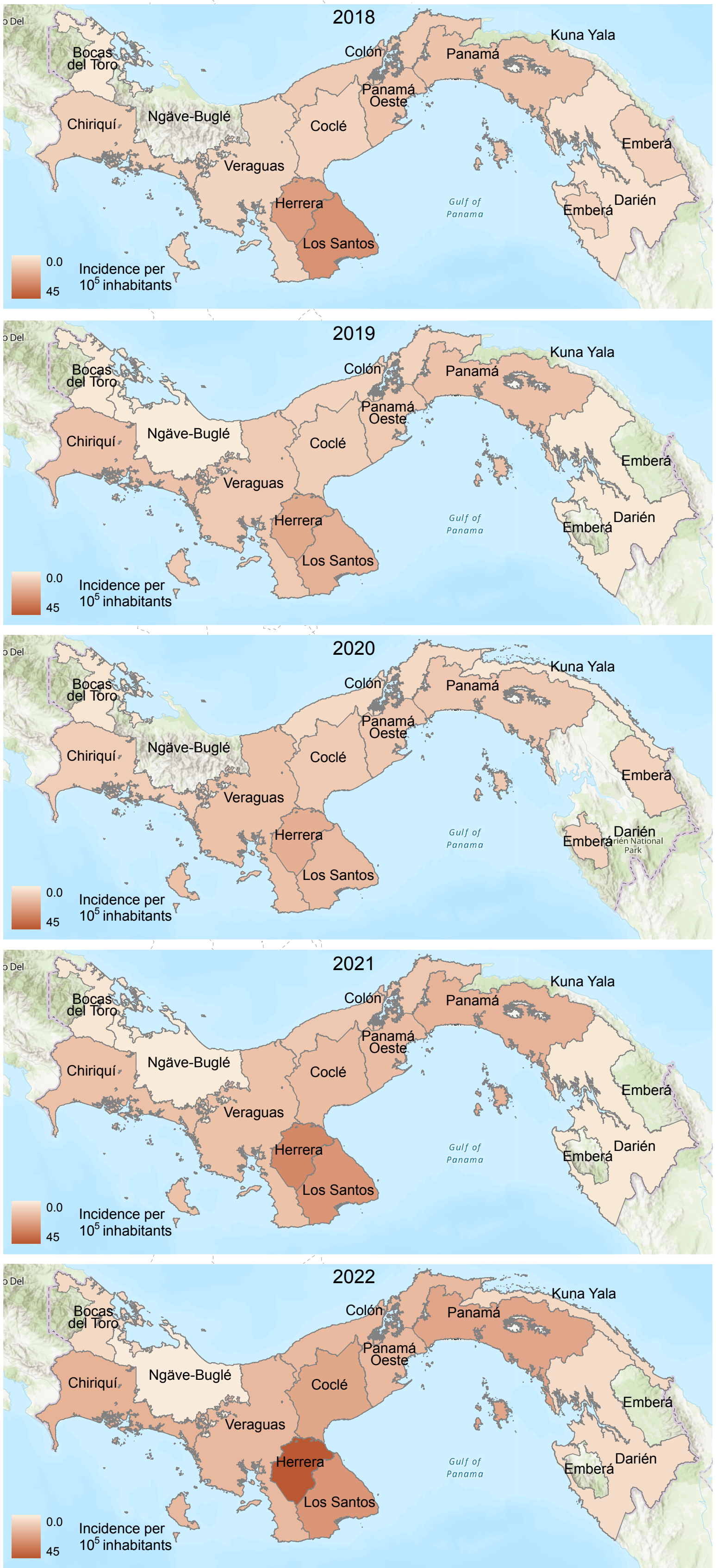

**Lung Cancer**

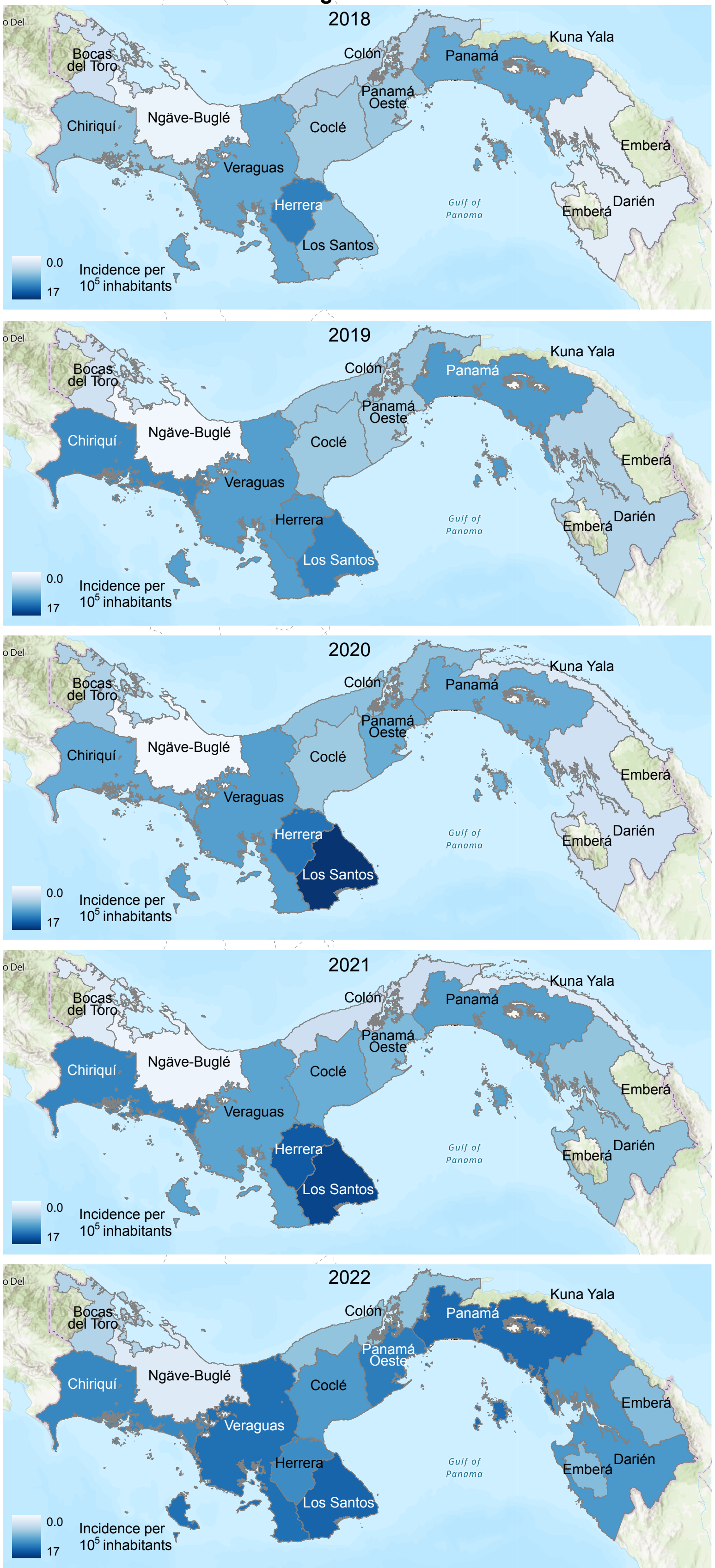
2018
2019
2020
2021
2022
Bocas del Toro
Chiriquí
Ngäve-Buglé
Veraguas
Herrera
Los Santos
Coclé
Colón
Panamá Oeste
Panamá
Kuna Yala
Emberá
Darién
Gulf of Panama
0.0
17
Incidence per $10^5$ inhabitants

### Skin Cancer

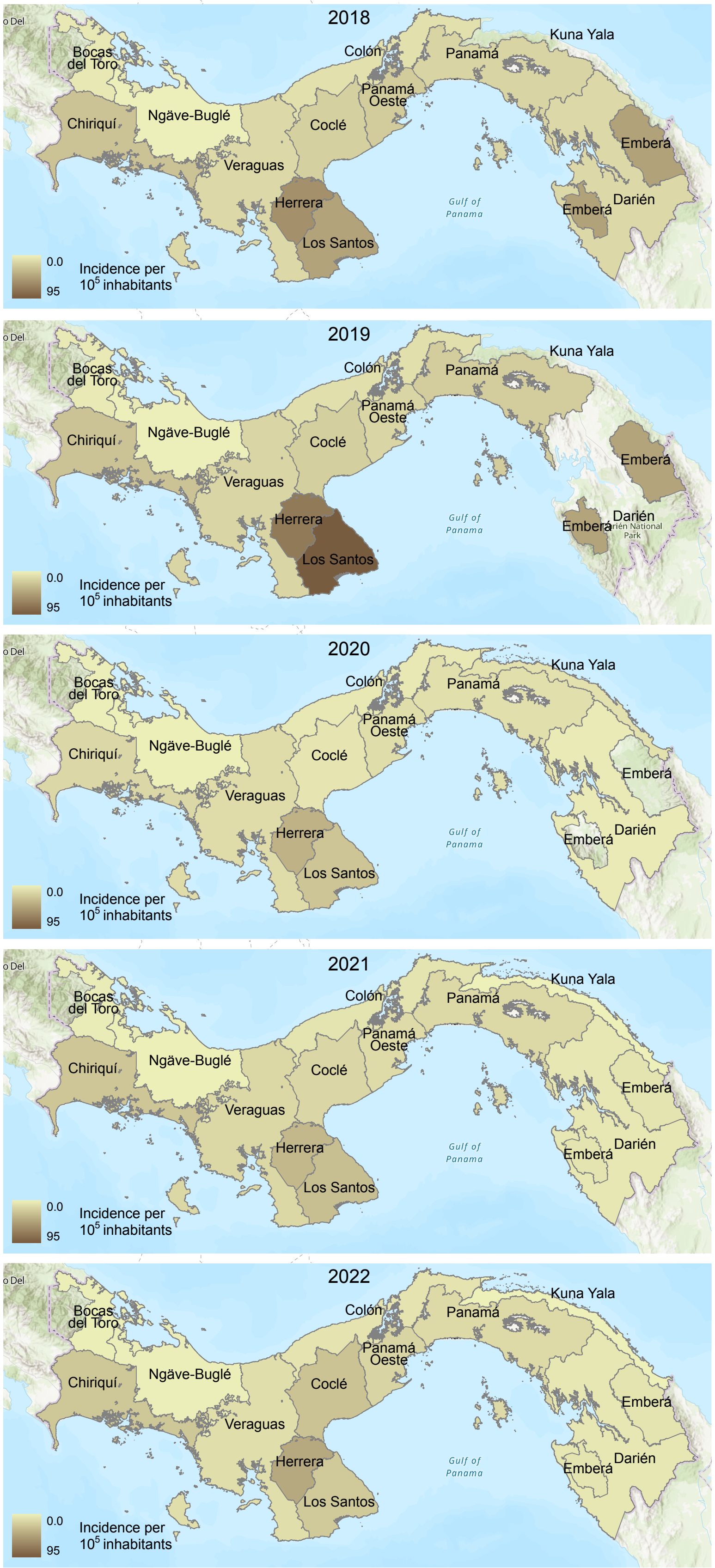
2018
2019
2020
2021
2022
Bocas del Toro
Chiriquí
Ngäve-Buglé
Veraguas
Coclé
Colón
Panamá Oeste
Panamá
Kuna Yala
Emberá
Darién
Herrera
Los Santos
Gulf of Panama
0.0
95
Incidence per $10^5$ inhabitants

**Breast Cancer**

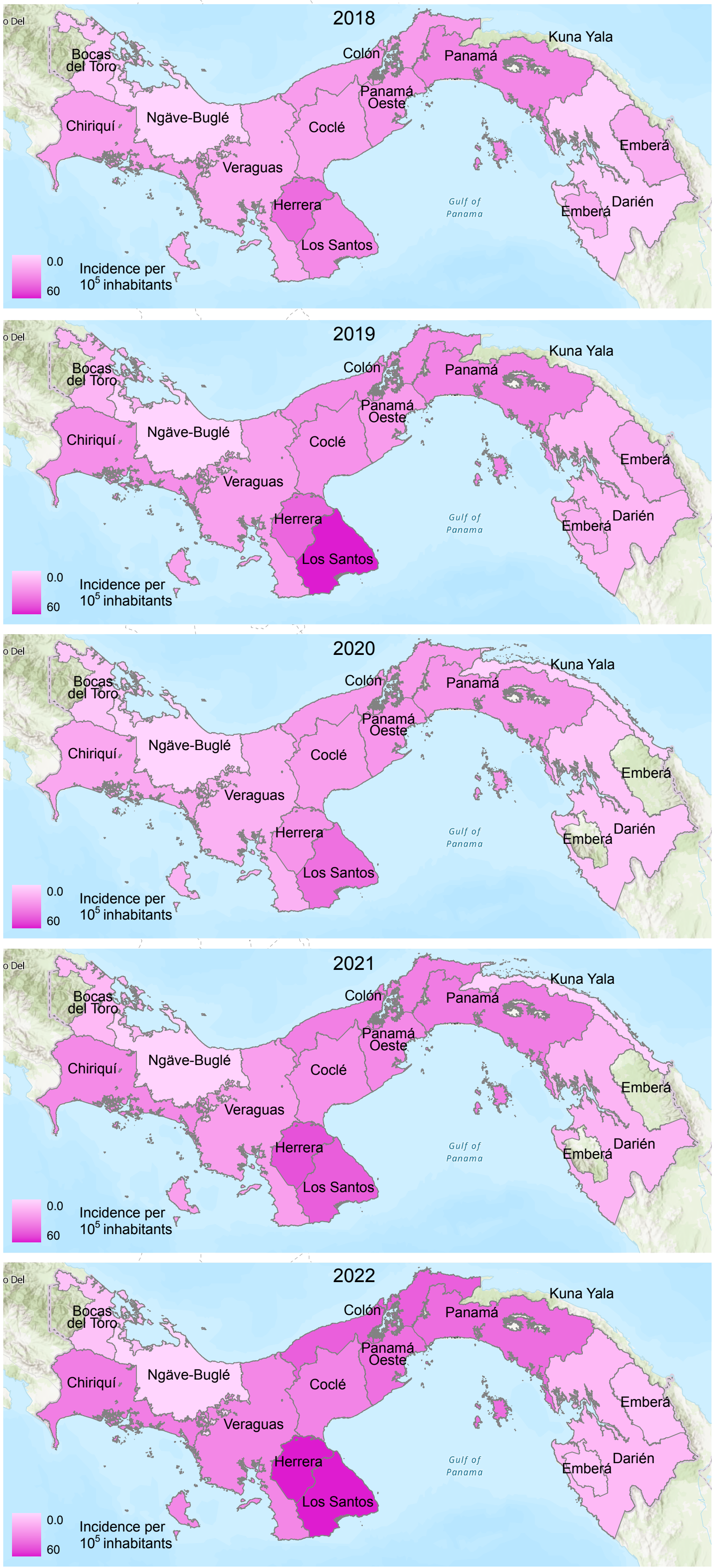

### Cervical Cancer

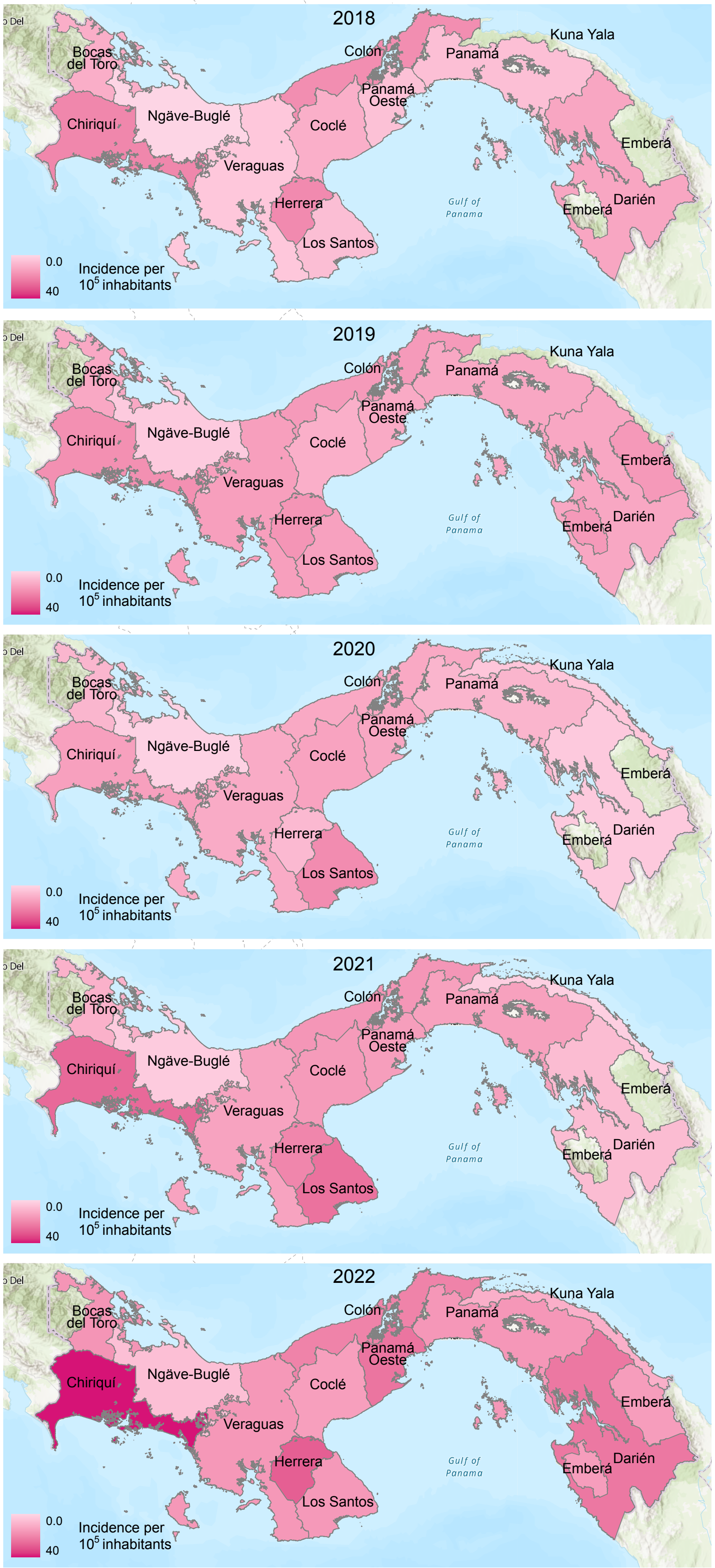

### Prostate Cancer

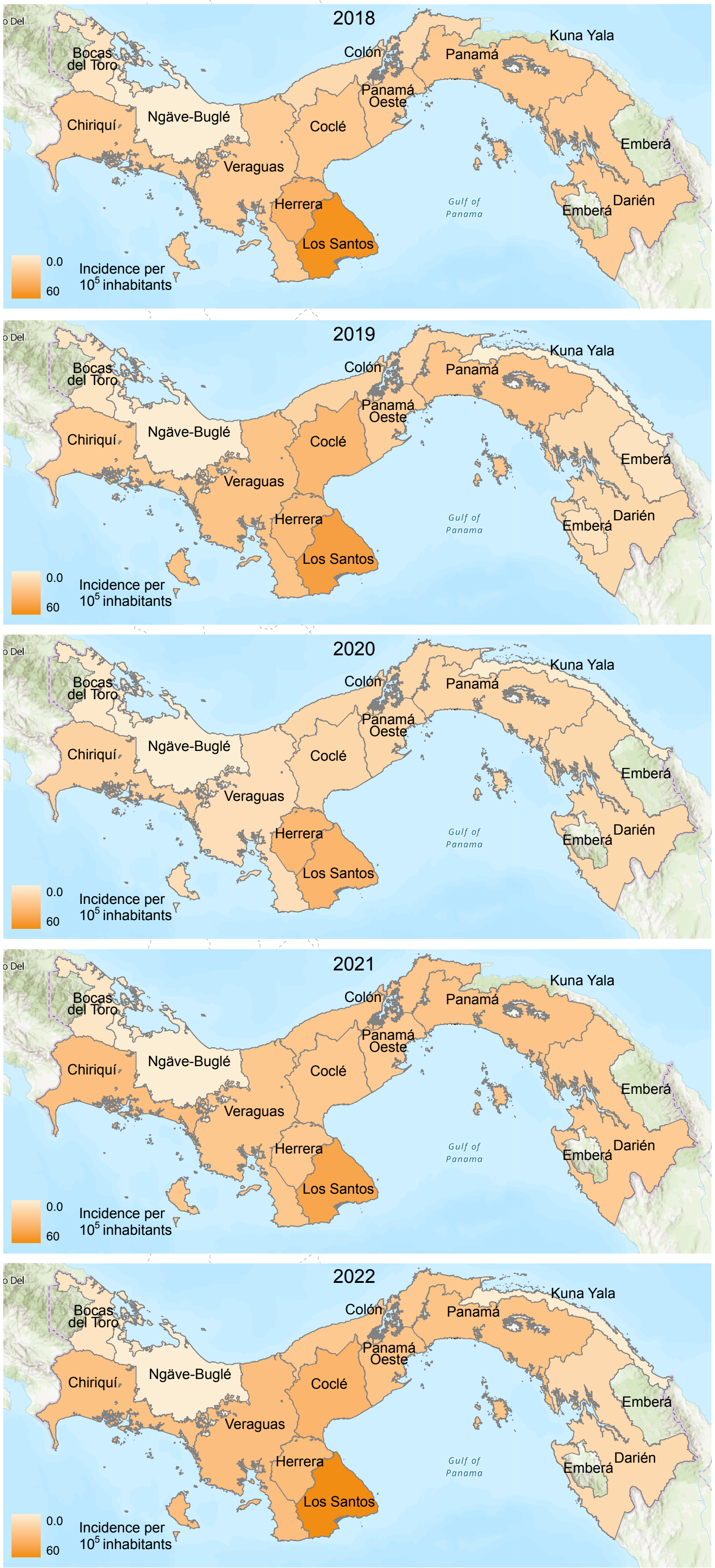
